# Factors associated with mental health disorders among students at Busitema University, an exploratory qualitative study among students at Mbale and Busia campus

**DOI:** 10.1101/2023.07.05.23291819

**Authors:** Enid Kawala Kagoya, Joseph.L. Mpagi, Paul Waako, Julius Wandabwa, Biira Saphina, Elizabeth Birabwa, Sophie Acon, Daniel Otim, Samuelbaker Kucel, Joseph Kirabira

**Affiliations:** Institute of Public Health, Department of Community Health, Busitema University, Faculty of Health Sciences, P.O Box 1460 Mbale, Uganda; Deans Office, Department of Academics, Research and Innovation, Busitema University, Faculty of Health Sciences, P.O Box 1460 Mbale, Uganda; Office vice chancellor, Busitema University, P.O Box 236, Busia, Uganda; Department of Psychiatry, Busitema University, Faculty of Health Sciences, P.O Box 1460 Mbale, Uganda; Department of Gender, Busitema University, P.O Box, P.O.BOX 236, Tororo; Dean of Students, Busitema University, P.O Box, P.O.BOX 236, Tororo

**Keywords:** Mental Health, Disorders

## Abstract

**Background:** Following the loss of several students at Busitema University faculty of health sciences and other branches, there was an urgent need to understand the factors contributing to the death of the students and it was anticipated that most of them succumbed to mental health issues. This study aimed to explore the potential factors associated with mental health disorders among students at Busitema University.

**Methods:** Key informant interviews were conducted among the students who were diagnosed with mental health disorders. A total of 42 key informant interviews were conducted following a well-structured interview grid. All participants consented during the study. Each interview was audiotaped and recordings were later subjected to verbatim transcription. Each transcript was carefully reviewed by the principal investigator prior to the analysis. Thematic content analysis was done following a deductive approach. Dedoose software was used to support the coding and categorization of thematic areas.

**Results:** The results indicate that several factors associated with mental health disorders included alcohol and substance use, poor learning environment, stringent and unfavourable university policies, the big gap that exists between students and administration, relationship challenges, academic pressure, family factors (Broken families, poverty), Gambling, lack of curricular activities, poor counselling services and political pressures.

**Conclusions:** Considering the high propensity of mental health issues that hinder the success of students at the university level. It’s worth paramount for universities to continually evaluate the mental health of their students and tailor treatment programs and other cost-effective interventions to specifically target students.

## Introduction

Mental health-related issues have been a matter of concern for quite a long time within universities and other institutions of learning yet they earn little or less attention, regard and interest from university administrators, the Uganda National Council for Higher Education as well as the Ministry of Education and Sports. Less emphasis has been put on students’ mental health and inadequate measures to ensure that the mental challenges of students are handled rightfully to achieve high levels of mental health [1]. The mental health of students at major universities is alarming and needs to be treated with much attention[2]. Majority of the undergraduate students experience high levels of mental illness which in the short and long run contributes to their poor performance and discomfort[3]. The majority of the population is battling with mental health issues including students at tertiary levels and other institutions of higher learning[4]. While some of the universities and schools decided to offer on-campus services like counselling sessions and mental health talks, some institutions have not designed such initiatives and many students are left without any help or solutions to the problems they face, in fact, most of the students who screen positive for some mental health disorders like depression and suicide do not receive help.

Most students with mental health disorders are likely to experience challenges like impaired cognitive function, drug and substance abuse, retarded / declined performance while at school and other anticipated learning disabilities., and learning disabilities[5]. Most of the anticipated drugs of use include alcohol and tobacco, some smoke cigarettes that in the short or long run affect their normal body functioning and some are associated with risky sexual behavior[6]. For this reason, therefore, there is a total affirmation that mental illnesses among university students are higher compared to people in other environments. Several factors have contributed to the mental distress and discomfort among students at universities like the learning /studying/ Living environment at the university, stringent policies at the university, Relationship challenges, academic stressors, the big gap that exists between the administration and the students, political pressures, gambling and above family factors have had a very significant contribution on the mental health status of students[7]. The prevalence of the conditions tends to be a notch higher among female students than male students. Universities have tight schedules and continuous sequences of study, which affects the student’s performance and their mental well-being[8]. Some of the predisposing factors are avoidable, while others are accustomed and tied to the students[9]. Therefore, it is prudent to come up with measures to ensure control and regulation of the inherent situation of undergraduate students, especially at higher institutions of learning. Mood disruptions are only one of the many mental health problems that undergraduate students face[8]. Although mental health specialists emphasize the need to talk about such concerns, students often regard these pressures as typical as the definition of academic lifestyle especially in medical schools in Uganda[1]. The majority of the students end up not getting help from the concerned parties because of their busy schedules at the wards and while monitoring their final year projects and pressure for exams as well. However, solving the problem is arguably easy by addressing some of the major health conditions that most undergraduate students experience.

Statistically, more than half of the students in public universities in Uganda suffer from depression and anxiety[10]. Similarly, a poll of undergraduate students at Mbarara University of Science and Technology in Western Uganda found that many students had suffered from mental health disorders such as anxiety and depression [11] while in the united kingdom,[12] showed that the prevalence of mental health disorders such as anxiety and depression cases are a notch higher in medical school compared to the general non-student community of the same age[8], which supports these findings. However, less has been done to ascertain and understand the factors associated with mental health disorders among undergraduate students in Uganda. Therefore the aim of the study was to explicitly explore the factors associated with mental health disorders among students at Busitema University’s faculty of health sciences and faculty of Engineering.

### Objectives

1. To explore factors associated with mental health disorders among students at Busitema University, as a case of the faculty of health sciences and Engineering.

### Study Site

The study was conducted at two compasses of Busitema University that is Busitema University Faculty of Health Sciences (Mbale Campus) and Busitema University Faculty of Engineering (Busitema main campus). Busitema University Faculty of Health Sciences is housed under Mbale Regional Referral Hospital in the Mbale district of Eastern Uganda which serves patients from 16 districts. It mainly offers medical courses at the undergraduate and postgraduate levels which include Bachelor in Medicine and Surgery, nursing, and Anesthesia. The second setting will be Busitema University, Faculty of Engineering which is located in Busitema, Tororo district, and mainly offers Engineering courses at diploma and bachelor’s levels such as mechanical and civil engineering among others.

## Materials and methods

### Study Designs

This was a descriptive qualitative study among students at two campuses of Busitema University a case of the faculty of health sciences and Engineering.

### Participants

The study basically targeted undergraduate students enrolled in Busitema University, either at the faculty of health sciences or the faculty of engineering who were diagnosed with one or more mental health disorders.

### Data Collection, instruments and Analysis

Key informant interviews were conducted among the students who were diagnosed with mental health disorders. A total of 42 key informant interviews were conducted following a well-structured interview grid. The interviews were conducted between 30^th^/March 2023 and 9^th^ April 2023 for Busia campus and 12^th^-20^th^/April/2023 for Mbale campus. All participants consented during the study. Each interview was audiotaped and recordings were later subjected to verbatim transcription. Each transcript was carefully reviewed by the principal investigator prior to the analysis. Thematic content analysis was done following a deductive approach. Dedoose software was used to support the coding and categorization of thematic areas.

### Ethical Consideration

Ethical approval was sought and obtained from the Research Ethics Committees (REC) of Busitema University, the faculty of Health Sciences BUFHS-2022-11 and the National Council for Science and Technology (UNSCT-Number: HS2700ES).

All interviews were conducted in safe and secure places with only one student at a time to ensure privacy and confidentiality of shared information. Administrative clearance was sought from the vice chancellor’s office to collect data from the students at both campuses. A written consent was obtained from all participants.

All methods and procedures were performed in accordance to all relevant national and international guidelines for conducting research involving human participants during COVID-19 pandemic.

For the sake of confidentiality, unique identifiers were given to all the participants with a number depending on the campus, course and year of study for example FHS/FOE 111 BNS

## Results

### Social demographics of participants

Social demographics of participants.
| Faculty | Male | Female | 1<sup>st</sup><br>year | 2<sup>nd</sup> year | 3<sup>rd</sup> year | 4<sup>th</sup> Year | 5<sup>th</sup> year |
| --- | --- | --- | --- | --- | --- | --- | --- |
| Health Sciences | 9 | 11 | 5 | 5 | 6 | 1 | 3 |
| Engineering | 14 | 6 | 6 | 4 | 6 | 6 |  |
| **Total** | **23** | **20** | **9** | **9** | **12** | **7** | **3** |

### Thematic Areas

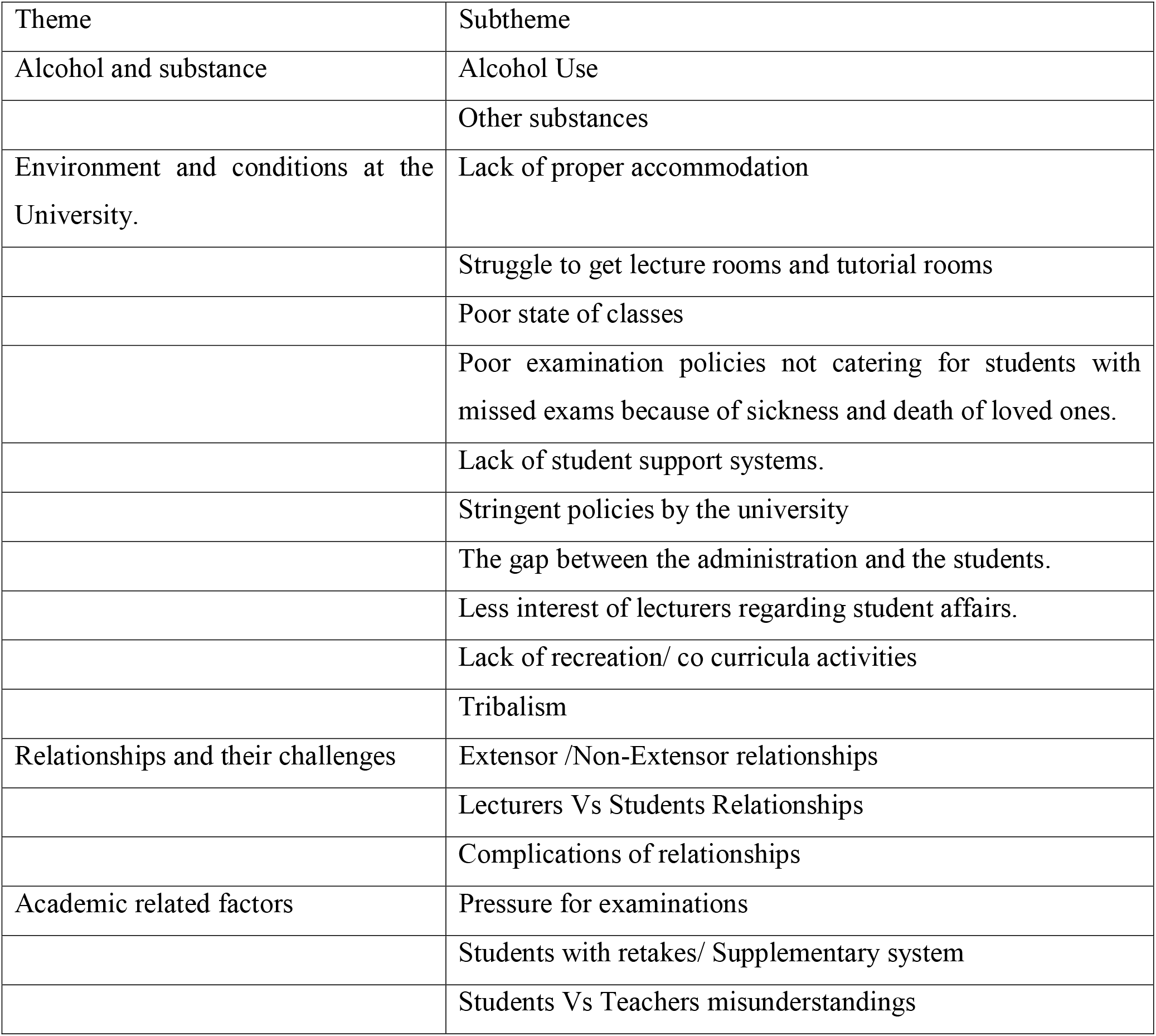
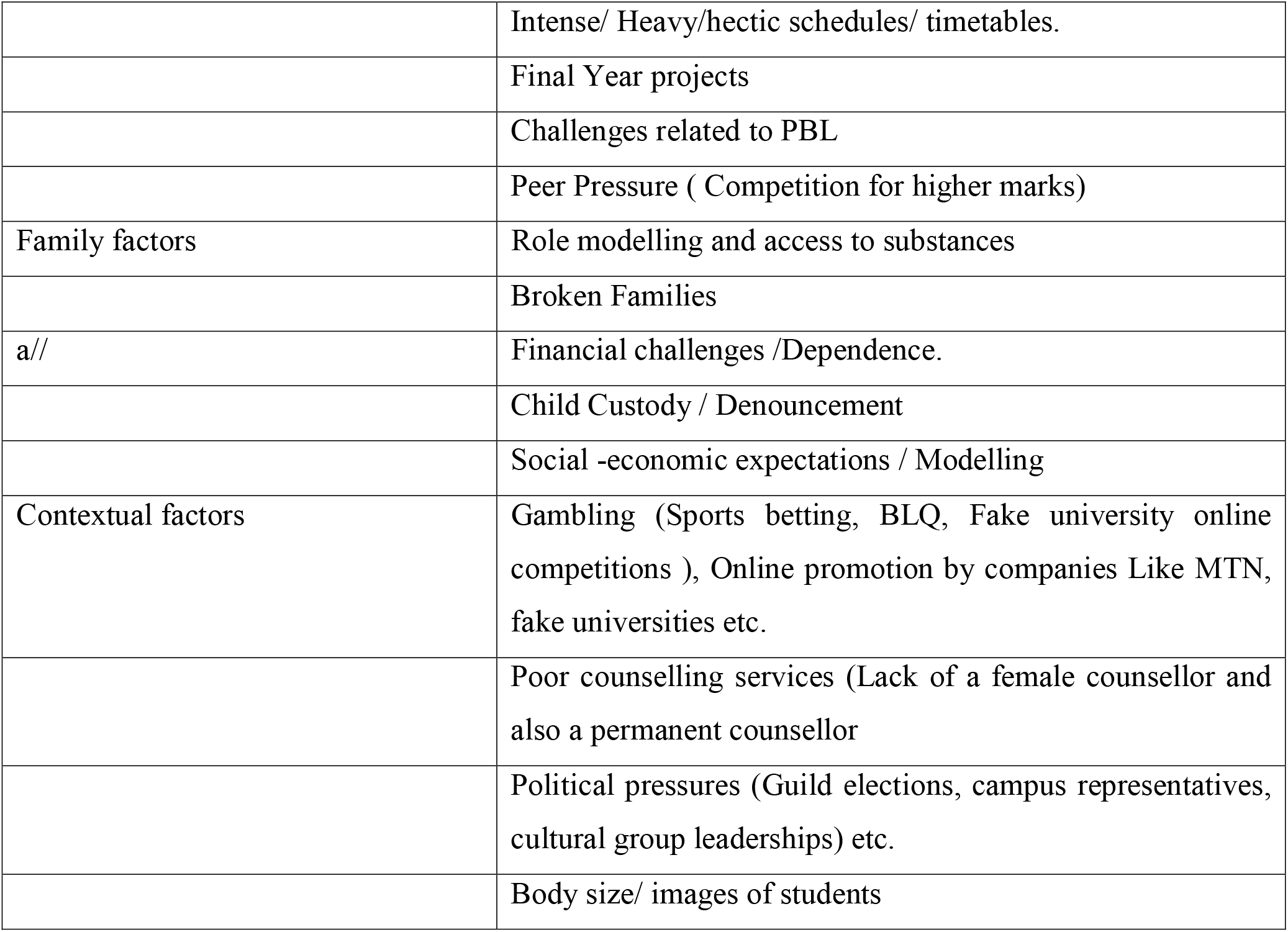

### Theme 1: Alcohol and substance use

Alcohol and recreational substances were commonly used by students at both campuses. Alcohol is the leading cause of many disorders and deaths for campus students, while some abuse drugs to induce their studying habits and boost sleep.

### Subtheme 1: Use of Alcohol

Some of the students at both campuses acknowledged that they used alcohol. The consumption of aslcohol was ignited by their appearance at social events like parties and the urge to have fun with their colleagues.

*“I take wines, Smirnoff, but once in a while, Like at social events like parties, when I get a chance to go to club etc. Just while socializing with friends and once in a while having fun. (**FHS, Male, 5, MBchB, 21-25 years)”***

*“I don’t take Malwa but I take Uganda Waragi because it works better for me, especially during the evening hours when I want to concentrate more and summarize all that I read the whole day **(HS, Male, BNA, 2, 25-30 years)**”*.

*“I take beer every weekend just to relieve must self from the stress. I take like 2 tuskers just to feel better after too much work at school, (**FE, Male, 3, AMI, 21-25 years)”***

Some acknowledged having taken beer but under influence of peers and curiosity while others reported having taken alcohol to calm down their frustrations and other personal challenges.

*“I have ever taken beer when we went for a party and I decided to take soda but everyone was looking at me and because almost all of them were taking beer, I felt out of place and decided to request a beer and it made me crazy, I promised not to take it again **(FE, Male, 4, WAR, 25-30 years,)**”*.

### Subtheme 2: Consumption of other substances

Besides alcohol, students also abuse marijuana, coffee, cocaine, ecstasy, and benzodiazepines. The dire need and desire to abuse drugs for various purported gains may lead to health complications resulting in death and body organ failure. Some students reported having used other substances like marijuana and local brew (Malwa). The consumption of the substances was a result of peer pressure and also anticipation that they would relieve them from the academic stress and pressures.

*“I used to use a lot of coffee and without it, I thought I would not function, feel sad in class and tired all the time. If I fail to get coffee, I would get a Coca-Cola or Pepsi because I knew they had caffeine and it would make me energetic though it brought me problems because I took it in high amounts and took me back to the depression” crying……….2 minutes **(HS, Female,5, MBchB, 22-30 years,)**”*.

*“It was one time when I tested marijuana because I had lost appetite, it was hard for me to sleep and I was so stressed but it didn’t do me well but I later stopped**(HS, Female, 5, MBchB, 25-30 years)**”*.

*“Some of us smoke a lot and take alcohol for example I know of a colleague who keeps crying when she is frustrated, often smokes weed and takes alcohol almost on a daily basis, he is actually the same person who give me some but I don’t do it on a daily (**HS, Male, MBchB, 4, 21-24 years)***.

### Theme 2: Environment and conditions at the University

Most of the students reported that the university did not have a conducive environment (social and academic) which predisposed them to most mental health disorders.

### Subtheme 1: Lack of proper accommodation and strict rules at the hostel

Some of the students referred to their hostels as unconducive for them due to their state and the unfavorable conditions attached to them. The final-year students as well as some students were not content with the rules related to vacating the hostel after the first year because it affected their security and also utilization of the faculty library.

***“****The living conditions at the hostel because I joined Compass expecting a life better than high school but life at the hostel was worse than my former high school. We were put up in a very small room, four of us, the bathrooms didn’t have water and then there was a day I had a running stomach, I had to fetch water downstairs for me to use the toilet, it was hard and some of our colleagues ended up getting blisters because of carrying water from a far distance from the hostels. The fact that we don’t have a better place to stay, traumatizes us. (**HS, Female, 5, MBchB, 25-30 years)”***.

*“We cannot use the library because when you get to the second year, you are told to vacate the premises to leave space for first years, if you are far from the Library, it’s hard for you to use it like those just next to it. (**HS, Male, 5, MBchB, 25-30 years)”***.

### Subtheme 2: Struggle to get lecture rooms and tutorial rooms

The struggle to get lecture rooms to students at the faculty of health sciences also predisposed them to mental health issues. The study indicated that there were few lecture rooms for use by students at the faculty of health sciences to the extent of studying in shifts (first years -3^rd^ years). Some ended up missing and postponing lectures and tutorials because of a lack of space.

*“I can imagine that sometimes we have to wait for the first years to finish their lectures then we start ours, so if they have a lecture for the whole day, then we end up not studying, I keep wondering why us …..(**HS, Female, 3, MBchB, 35-40 years)”***.

*“Sometimes we have exams at the same time and our class is big, we can’t fit at the hostel, we end up sitting on verandas and trinity which is ever smelling, (**HS, Male, 1, BNS, 21-24 years)”***.

### Subtheme 3: Poor state of classes especially at the faculty of health sciences

The study indicated that most of the students were not comfortable with the classroom environment at the faculty of health sciences which made them feel like they chose the wrong university. Some of the students reported that the lecture room was leaking and with only one bulb. They also informed us that due to the large numbers, some of them sit out of class during examinations and while they were writing their papers, they are showered with rain and papers got wet.

*“When I joined the university, I expected to see buildings like those at the main compass but I was shocked especially when I was sitting for my first paper on Ethics, all of us who were seated at the front of the shade got wet:……cry: The lecture also got wet …ohhhhhhhh my God….i felt like dropping out of the course and waiting to apply elsewhere***, (*HS, Male, 1, BNA, 21-24 years)”***

### Subtheme 4: Lack of student support systems, Stringent and unfavorable university policies

The study indicated that the university did not have favourable examination policies, especially for the students who missed exams due to certain reasons like the death of a loved one, there was no affirmative action at the university which affected most of the students who ended up either giving up on the course or take dead years for them get relieved.

*“The university is so tight or hard on students as they don’t care for students who aren’t healthy as in sick or after losing a loved one. It’s hard for the students to do missed papers just because of just loss of a loved one, even when you had completed your rotation, you have to rotate again, it’s just difficult, (**HS, Male, BNS,4, 21-24 years.)***

### Subtheme 5: The gap between the students and the administration

The study found that there was a gap between students and the administration more so with students in the faculty of engineering. It was hard for them to share their mental health-related challenges with the administration and staff as well and instead resorted to informing the faculty leadership who had no solutions to their problems. Staff and administration had less or no time to attend student-related activities which also did not please students.

*“The administration is far away from the students so the students should be each other’s keepers (**FE, Male, 4, BCT, 21-24 years)”***

*“They fear contacting the administration because there is a big gap between students and the administration and they don’t want to attend sessions of that nature to support to students (**FE, Female, 4, WAR, 25-30 years)”***.

### Subtheme 6: Tribalism

The study also identified tribalism as one of the factors which contributed to mental health disorders among students at the university. Some students acknowledged that tribalism was a vice eating up the university and affecting the performance of students at the two compasses. It was reported that lecturers from some tribes favored their tribe mates to the level of organizing separate sessions with them or for them. The study also identified a few aspects related to scoring their tribe mates highly in the courses taught but some lecturers. Students who were not favored in that regard felt left out and their mental health was affected.

*“We have a lecturer (name disclosed), during his lecture, if you are not his tribe mate, all your answers seem to be wrong to him, we keep wondering whether there are answers associated with tribe, (**FE, Male, 1, DGE, 21-24 years)”***

*“One of the best departments at this faculty has a lecturer who favors his tribe mates, even during sessions, we have observed it, I cannot forge the time he called them for a separate session and set the same things he taught them, anyway, we got to know and that traumatized some of us… (**HS, Male, 4, MBchB,25-30 years)”***

*“...........have you realized that there are some course units passed by students of some tribes, some of us didn’t bother reading hard because we know it for those guys…. (**HS, Male, 5, MB, 25-30 years)”***

### Subtheme 7: Lack of recreation/ co-curricular activities

Unlike students at the faculty of engineering, students at the faculty of health sciences reported having no time and space for co-curricular activities at their compass. This was related to lack of space as they didn’t have grounds or fields for the curricular activities, curricular activities were not in any way incorporated in their timetables and students were made buy with exams, ward work and others which made them forget about such activities.

*“We don’t have a timetable for co-curricular activities at this faculty unlike other faculties (**HS, Male, 1, BNA, 31-35 years)”***

*“Some of us have talents which we can’t put to use, we recently created a group and days where students could come and showcase their talents but it was an initiative for us fifth years and now that we are leaving, I think we are leaving with it (**HS, Male, 5, MBchB, 25-30 years)”***

### Subtheme 8: Students VS students misunderstandings

The study also acknowledged that there were some misunderstandings between lecturers and students which also contributed to their mental unwell being. The misunderstandings culminated form arguments during class and tutorial sessions.

*“There was a time in 3^rd^ year when we were doing CVS anatomy, so we were told to take out our bags to the lower block and return for the exam on return, the lecturer told us not to sit for the paper and go ahead to sit of it when next offered. (**HS, Female, 5, MBchB, 21-24 years)***.

#### So what happened that after?

*“We had to plead for him but he refused. We had to just believe that we had a retake and move on but depressed me for a while. We were just good children, didn’t break rules, and were not defiant, but that is how he punished us. (**FHS, Female, 5, MBchB, 21-24 years)***.

#### Did it affect your mental health?

*“It was so painful and all of us cried, He gave us another paper after 2 weeks but it was very hard OMG, he instead gave us a theory yet we were meant to do a practical, it was traumatizing. He gave us another paper after 2 weeks but it was very hard OMG, he instead gave us a theory yet we were meant to do a practical, it was traumatizing. We did not pass very well, almost all of us got Cs yet some of us want to specialize in cardiology and you can’t do it with a C. (**HS, Female, 5, MBchB, 21-24 years)”***.

*“There was an argument in class on leadership and management during a tutorial report back with one of the tutors we had an argument of who is a leader and who is the manager. People were saying that a leader is above a manager, I gave in my opinion but we did not come to a conclusion and I and the tutor had an argument we didn’t agree and he was pissed. At the end of the report back, he allowed me to talk and asked me do you know that you have a probe and also asked me” are you willing to be helped”, he responded and told me to visit a counsellor. I there and then asked the class representative to give me the contact for the counsellor and counselled me and gave me the assignment to read a book about “River of life **(HS, 1, BNA, 30-35 years)”***.

### Subtheme 9: Expectations of Lecturers from Students

Some of the students acknowledged that some lecturers or teachers were too hard on them to the extent of embarrassing them during the lecture sessions especially when they seemed to know what the expectations of the teachers were. The study showed that some teachers go to class expecting students to have read ahead of them and thus expect students to know some basics however, lecturers kept embarrassing those who did not read ahead of the lecture or who were unable to answer their questions.

*“Some lectures embarrass us sometimes in class because they expect us to know everything and consider it to be a crime not to know something. We used not to show that we had mental health issues but some of our colleagues were going through a lot so lecturers should style up (**FE, Female, 2, BEE, 25-30 years)**”*

### Theme 2: Relationships and challenges associated to them

The study also found that there were relationship issues among students (students vs. students) and (students vs. lecturers) and their associated consequences or complications. The relationships in the long run or short term also contributed to the mentally unwell being of students at both campuses.

### Subtheme 1: Extensor /non-extensor relationships

The study also identified some relations ships between students (extensors vs non-extensors). Some of the students acknowledged having been in such relationships which were productive though they turned into non-productive after some time. The unproductivity was associated with first-time exploration and loss of interest in the students and others were related to the ushering in of first years who were “young and beautiful” and hence abandoning the previous relationships. Those who were abandoned were left in the grey lights of a cold night hence affecting their mental health.

*“I had a boyfriend and things did not go well with us. We were okay at first but we broke up. He was a student at Busitema, faculty of health sciences but I thought he was not worth my time. He could not value me and could only care when he wanted something from me. But when we broke up, we abused me terribly which was not good. I had had sex with him twice to bind us together (**HS, Female, 2, MBchB, 21-24 years)***.

*I was in a relationship with a student leader, the guy had money and was very caring and supportive but when the first-year joined, he abandoned me and got a fresher doing water resources engineering…..of course, I was heartbroken and missed classes for a week. (**FE, Female, 3, WAR, 21-24 years)***.

### Subtheme 2: Lecturer/Students relationships

Lecturer vs student relationships were one of the factors that contributed to the mentally unwell being of the students at both compasses. Some of the students acknowledged having engaged in serious romantic relationships with lecturers however, the once enjoyable relationships turned into a devil hug in the dawn leaving them in a state of quagmire and mental health distortion,

*“There were relationship issues that happened while in my first year, I was in love with this lecturer who promised me heaven on earth …..Crying….I gave in to his feelings but later I heard that he had another lover, it took me a lot of time to recover because it kept running in my mind (**HS, Female, 3, MBchB, 21-24 years)**”*.

*“I was in a relationship with a lecturer (name not mentioned), he approached me when I was in year 2, appreciated me and used to do a lot for me. I had sex with him like four times and after some time, he trashed me and started abusing me and talking about my weight, and body shape and told me to give him a break. Awwwwwww (cried…………….). My friend who knew about our relationship used to laugh at me when the guy dumped me, it led to another phase of depression (**HS, Female, 3, MBchB, 21-24 years)**”*

### Subtheme 3: Complications of relationship

The study indicated that complications of relations also affected the mental well-being of the students at both campuses. The complications identified were related to abortions, sexually transmitted diseases, and heartbreaks to mention but a few.

“During my relationship with some lecturer, I think the spouse used to note the times I used to call him, so one day the wife gives me a call and we plan to meet, I didn’t know she was the wife to Dr …..the lady embarrassed me in public, slapped me and tore my clothes. (**HS, Female, 1, MBchB, 25-30 years)**”

*“We are struggling with STDs and abortions which makes us worried and sick all the time. There are a lot of unwanted pregnancies and abortions among students (**FE, Male, 4, MB, 21-24 years)**”*

*“Issues of relationships, as you know am of age, and I keep asking myself when am going to get married I had a relationship before but the person just wanted to use me all the time and want to take me anywhere. But my worry was lack of a potential person to love me etc. (**HS, Female, 3, BNS, 25-30 years)**”*.

### Theme 3: Academic pressures

The pressure attached to the academic programs at both campuses was a factor of consideration in relation to the mental well-being of students. Students at both compasses reported that they did not have time to do anything that was non-academic related. Besides that teachers were not teaching them how to balance life and work. The doctrines at both campuses were bout reading, excelling etc. which was accompanied by heavy schedules, and too many tests and exams and it was worse with the faculty of health sciences where the PBL system seemed more hectic.

### Sub Theme 1: Pressure for exams

Pressure for examinations was one of the factors that raised the eyebrows of most students at both compasses. The too much reading during the time of examinations and the less confidence and assurance of passing created more pressure leading to mental health disorders like panic attacks, depression and also the use of alcohol and other substances.

*“Academic issues, where you expect us to do much but we do less, for example, there are some course units which are demanding, you have to do tutorials and then report back and lectures are intense but it’s the system, we have nothing to do about it**. (HS, Female, 3, BNS, 21-24 years)**”*.

*“Academic pressure where we have to read when you are hungry, the timetables were too tight and I had no money, I had so many issues at the same time **(HS, Male, 3, BNA, 21-24 years)**”*

*“It was academic stress, it was after a viva, I thought I knew and I had read but when I went for the viva, I felt humiliated by the fact that the person who was conducting the viva didn’t give me a chance to express myself very well, **(HS, Female, 3, BNS, 21-24 years)**”l*.

### Subtheme 2: Students with retakes

Regarding the problem of retakes, some of the students acknowledged having gotten retakes which disempowered them academically and affected their mental well-being. Some of them felt like retakes were another factor contributing to the withdrawal of students from some courses because the lecturer kept seeing them as failures before other students which led to depression.

*“There is also a bias for students with retakes which makes them keep getting retakes so setting up a support system for them is better than tormenting them and using the sarcastic word during lectures most of them get depressed and change because of a retake. The moment they get a lot of retakes, they are reduced and they lose hope and withdraw. (**HS, Female, 5, MBchB, 21-24 years)***.

*“I was also depressed when I got my worst mark which was a D+ which made me change my lifestyle and withdraw from leadership roles **(FE, Male, 2, AMI, 21-24 years)***.

The fear of retakes also contributed to anxiety among the students and the desire to pass highly which affected their normal body functioning and psychological well-being.

*“For anxiety, I was worried about retakes, assignments, and the desire to pass highly and want to be smart. When I went for clinical years, my anxiety went up, I had spiking blood pressure and the anxiety was too much. **(HS, Female, 5, MBchB, 21 -24 years)***

### Subtheme 3: Intense/ heavy schedules

The study revealed that the schedules at the compasses were intense and heavy not giving them time to relax and engage in personal work for self-development. The study also revealed that some of the students at both camp passes were extensors who had to balance between work and school without study leave.

*”I was working, and then I had school which had come around and then I had pressure from school, from work, from family, yeah concoctions of a lot (**HS, Male, 2, BNS, 25-30 years)”***.

*“Programs are so packed not give us time to refresh because we have a lot of tests and exams which drain us a lot (**FE, Female, BCT, 21-24 years)”***.

*“We haven’t been taught about life after medical school, it’s about life at medical school yet there is life after that (**HS, Male, 3, MBchB, 21-24 years)”***.

### Subtheme 4: PBL challenges

The study also found that Problem-Based Learning (PBL) was a new method of teaching and learning to some students at the faculty of health sciences. Some students reported having not used the PBL system before and therefore found it very demanding because it had a lot of tutorials which ii needed intense reading and understanding of the problems coupled with lectures and seminars. They felt drained at the end of the day.

*“PBL can drain, I can’t imagine the number of assignments I have to do in a day, yet we also have dissection sessions, lectures, discussions, etc, by the end of the day, you seem very tired with a migraine. (**HS, Male,1, BNA, 31-35 years)***.

*“When I had just joined Compass, I thought we were going to study the normal way but I found new things like tutorials, and SDLs which I was not used to **(HS, Female, 3, MBchB, 21-24*** **years).**

*“I have siblings at Compass who study for two hours maximum of four hours and their day is done but here, from class to presentation, to projects, heeeeeee it’s tiresome. (**FE, Male, 3, MEB, 21-24 years)***.

### Subtheme 5: Final Year projects

Results indicate that students at the faculty of engineering were struggling with their final year projects as most of them found them demanding in terms of time and financial resources. It was also discovered that they were not getting adequate support from their mentors or supervisors, especially in monitoring and evaluating the progress of their projects. This was attributed still on the big gap that exists between the teachers and the students. The failure to provide timely responses affected the mental well-being of students.

*“My thoughts were related to how I will finish the course and the final year project. The supervisors were not responding I had fear of being before the viva panel **FE, Female, 4, WAR, 25-30 years)”***.

*“Final year projects where supervisors are not supportive of us, they leave us to suffer alone and when you don’t do the right things, they are mad at us, (**FE, Male, 4, BEE, 25-30 years)***.

*“The loan scheme, it came in late around December and yet the course had started, so I was trying to balance between travelling from work to school even now they are back and forth movements. (**HS, Male, 1, MBchB, 25-30 years)***.

### Theme 4: Family associate factors

The study also identifies some family-associated factors which include the existence of broken families, family rejection, poverty and financial challenges, and child custody among others which left some of the students torn between school and home.

### Subtheme 1: Role modelling and access to some substances

The study informed us that some of the students were victims of alcohol and substance use which was highly driven by their family members like aunties, fathers and uncles. A few students informed us that they decided to engage in substance use because they used to see their parents take them at home and during parties. Others acknowledged having come from families where brewing was an economic activity, therefore, getting used to taking it because they had to taste the end product before it could access the market.

*“At home, I stay with my relative who makes Malwa so whenever we needed to take it to the local bars, she used to tell me to taste it that’s how I got used to taking Malwa and its sweet compared to beer, **(FE, Male, 2, DAG, 21-24 years)”***.

*“Everyone takes alcohol and shisha in our family, so if you come there and you don’t use them, you feel out of place. **(HS, Male, 3, MBchB, 21-24 years)”***.

### Subtheme 2: Broken families

The study also revealed some of the students were grappling with broken families making life both at school and home hard. Some students acknowledged that they were victims of gender-based violence witnessed at home by their parents, and some families had speared where most of them were just staying and living with their mothers as the sole bread winners in the family.

*“Iam not sure what time it was but I have had a hard time. It started in my childhood but I was a little better when I joined compass though it has affected me up to know. Our parents, just left us there. My parent was the first to leave after selling off everything and left us without a relative, young as we were. **(HS, Female, 2, MBchB, 21-24years)***.

### Why did your dad leave?

*I don’t know what happened to him but sold all things that we owned like land and then the rich guy who bought the land came and put down our plantations and started planting a lot of grass for his cows and we had nowhere to go so our relative took us to our other relatives in the village and life there was not good because our relatives had conflicts and assured us that were we were not their children (Takes a deep breath…..), we did not have food and other basic needs and even when we got sick, none bothered. **(HS, Female, 2, MBchB, 21-24years)***.

Some of the students were also abandoned by their parents which left them in child-headed families or with nowhere to go and they had to struggle to earn a living as well as meet the need of their siblings. This made their life at school and home hard because some of them were equally young to take on such gender roles and responsibilities.

*“After that, our relative also left ….I think because we are a burden to her, we were three (she cried for 3 minutes) and left me with my sibling and went with my little siblings and the relative in the village had also gone… so we were left there with nowhere to go, no one to be with us (shehehheh crying…) we kept depending on a relative for food. I was not going to school that time. We went to my other relative but she also chased us way but later they took us to some relative who treated us well up to now. **(HS, Female, 3, MBchB, 21-24 years)”***.

Gender-based violence at the family level was also a problem that affected the psychological well-being of some students. The study showed that some of the student’s feared going back home after their end-of-semester exams and key public holidays like Christmas because of the fights that existed between their parents.

*“I feared to go back home and I felt I needed not to be there and decided to keep in Mbale even when we have holidays because I know the community at home, people would laugh at me because my parent was involved in a certain act that killed her image and didn’t want to associate myself to such an act **(HS, Female, 4, BNA, 21-24 years)”***.

*“Whenever my parent comes home, he beats up my other parent and he comes when he is drunk, takes all the money from her leaving us not happy, there is day he beat her and took all the money she was meant to pay for my tuition, I feared to report that semester without tuition paid, **(FE, Female, 4, MEB, 21-24 years)***.

“*I had a family issue. I was disturbed by an incident that happened between my parents. There was a misunderstanding between my parents and spanned from things related to infidelity that made me depressed coupled with things that followed after that when I felt like I wanted to be independent and felt like I didn’t want to get attached to them anymore and didn’t want to talk to them, I was in good terms with one of them but because of that I hated the other but later ended up being with her which I thought could land me into problems. I felt like I want to be financially independent of them so I ended up getting involved in all kinds of business in Mbale because I even didn’t want to go back home **(HS, Male, 2, BNA, 25-30 years)”***.

### Subtheme 3: Financial-related factors

The study also revealed that financial-related factors greatly contributed to the mental well-being of students at the university. Most of the students were struggling financially, especially those from matrilineal families, the extensors were also grappling with work and those who had businesses as a source of funding also had some challenges as their businesses were not performing well due to competition and the nature of the economy at that time. Some students acknowledged having come from families that were not well-to-do with many dependents.

*“Home issues like financial issues because I am not a government-sponsored student and my parents have to pay it on time. My parents work but the defendants are many which makes tuition delayed and yet sometimes we are required to register in time and you keep thinking about that all the time, **(FE, Male, 1, BCT, 21-24 years)”***

*“It was when they sent us the government allowance and my money was stolen. My phone was hacked and over 4.6 million was stolen. I tried to settle a lot of issues like my rent, I had a lot of pressure from some people I had promised some money at home. My parent abused me and blamed me for not sending the money immediately to him. DW tried helping me but nothing was helping. We were doing GIT and I was stressed and that also increased the stress. But by the time we started doing head and neck, I had stabilized a little **(HS, Female, 2, BNS, 21-24 years)***

*“I started a small business but didn’t work out and tried all alternatives that would bring me money because I was a government student and none was paying for me tuition. All the government allowances were spent or wasted in such attempts to stat small business and wanting to be independent **(FE, Male, 3, WAR, 21-24 years)”***.

*“I was not on loan scheme, and then at one point was worried on how am going to take care of my finances, yeah am like the second born in the family, so I have other siblings who could be in need of the parents support, so I didn’t see anything working out for me*.

***FE, Male, 2, BCT, 31-34 years)*.**

### Subtheme 2: Child custody/denouncement

The student also found out that one of the factors that contributed to mental health disorders among students was child custody and denouncement. Some of the students were left to act as parents while as others were rejected as a result of a change of religion and not doing the expected courses of parents’ interests.

*“As you are aware my name and religious affiliation. I was born a Muslim but as I grew up, I decided to change religion and my parents rejected me up now. **(HS, Female, 1, MBchB, 25-27 years)”***

*“My burden has been taking care of my siblings, those who came after me of which I even tried, because my follower is a nurse, God has helped him he has got a job with the government. And then there is one graduating this year, think next week, then the three are in secondary level, one is going to senior six, one to senior five then the last one has joined senior two. The pain that comes to me …..is seeing the burden that has been left behind me, like pushing on with those that are in the lower classes but also could not be the case coz there is no one who has stepped into my shoes to cover up the gap. Personally I couldn’t continue to work for all those years. I also thought of upgrading, so now right now my challenge is where I am going to get the school fees. **(HS, Female, 3, BNS, 25-30 years)”***

### Theme 5: Contextual factors associated with mental health disorders among students

The study revealed that there were other contextual factors associated with mental health disorders among students at Busitema University. They included gambling like getting involved in sports betting, online competitions, lack of counselling services at the university, the body images of some students, social expectations, and social media to mention but a few.

### Subtheme 1: Gambling (Sports betting, BLQ, Fake university online competitions), Online promotion by companies Like MTN, fake universities etc

The study found out that gambling was one of the vices eating up the time and finances of students at both campuses. Some students acknowledged having engaged in some forms of gambling like betting especially when they had the “BLQ” betting competitions where they had to give in some money in anticipation of higher returns but instead lost. Some of them acknowledged having participated in online scholarship competitions where they wasted their money paying admission fees and application fees to the wrong universities (it was a smarm).

*“What led to my mental health issues was betting, I used to bet a lot and waste all my weekends betting. I also used to spend my money in betting and ended up lacking food to eat. The anxiety related to betting could make me lose my appetite and sleep and even concentration **(HS, Male, 2, BNA, 15-20 years)”***.

The struggle to get money prompted them to apply to international universities for scholarships but all this did not work out after realizing that it was a scam.

*“Considering all the pressures I had, I came across a link for UNICAF and some other international universities where they were giving scholarships to undergraduates, all most all my classmates applied, so I also got money and I applied but none of us got it because it was a fake call **(FE, Male, 3, BCT, 25-30 years)”***.

### Subtheme 2: Body size/ images of students

The study found out that some of the students were psychologically tortured because of their feelings and compliments about their body size and shape. It was worse during the lecture when they had to talk about obesity and its associated factors, fellow students kept pointing fingers at them which affected their mental well-being.

*“I also had an issue with body size or image which psychologically tortured me. All my classmates were small and I thought I was very big and need to become a little smaller or medium size, mine was kinda genetic but people would keep telling me to watch out and work out. (**HS, Female, 5, MBchB, 25-30 years)”***

***“****Then when we are in class while talking about the causes of some diseases, we would begin with the definition, causes, epidemiology etc and all that would come through was obesity and overweight and students would point figures at me and it used to traumatize me**(HS, Female, MBchB, 5, 21-24 years)”***

*“I had to go for extremely large bras, oversize and yet others were small, I used to wear sweaters all the time and used to cry all the time and I had a feeling of looking at myself in a mirror when I joined campus, I mixed it with academic issues and it worsened everything because it ended up getting low points, I had to come to Busitema yet I desired to join another university**. (FHS, Female, 5, MBchB, 21-24 years)”***

### Subtheme 3: Poor counselling services (Lack of a female counsellor and also a permanent counsellor

*“We don’t have counsellors here at the main compass, we end up telling the faculty dean or RCC who don’t know how to council and we can’t tell them everything (**FE, Male, 4, AMI, 25-30 years)”***.

*“We only have a male who is not there all the time and nothing like a female counsellor because some male counsellors don’t understand the problems of female students, for example, I can’t open up to a male counsellor about my relationship issues, and I feel shy(**HS, Female, 3, MB, 21-24 years)”***.

### Subtheme 4: Political pressures (Guild elections, campus representatives, cultural group leaderships) etc

Political ambitions and the associated pressures triggered some level of mental well-being. The anxiety associated with keeping in the race and successfully taking the position would throw a cold ice cube on the nerves of some students.

*“It was a political race and I was not financially not stable and I had no one to support. It put me into a state of depression and I used not to go for the parties to which they used to call me and I used to pray to God (**FE, Male, 1, BCT, 31-35 years)”***

*“My sibling was also traumatized because he did not have a job and yet he is the one to support us, he failed to get a job and he is now suffering, it also traumatizes me a lot. (**HS, Male, 3, BNA, 21-24 years)”***

### Subtheme 5: Social academic expectations/modelling

The study also revealed that parents as well as lecturers didn’t put into mind the aspect of balancing life with work or school which has drained some of the students. They struggled to balance school and social life which in the long run made them finish semesters without associating with people outside of the school space.

*“The expectations of being a medical student are high, I struggle to balance between life and school and have no other work beyond class cried for some seconds………but it stresses us, (**HS, Male, 5, MBchB, 21-24 years)”***.

*“Lecturers and parents as well should teach us how to balance life and work because medical school is bout work, no other life. You are always thinking about, class assignments, tutorials, what I know and what I don’t know etc, (**HS, Male, 1, BNA, 25-30 years)”***.

### Subtheme 8: Social Media and its side effects

Results indicated that social media was one of the associated factors with mental health disorders among students. Some students acknowledged that WhatsApp was the easiest mode of communication came along with so many messages that used to disorganize them during the day. Some of them informed us that watching TikTok videos with bad news, and nasty messages also traumatized them.

*“I think anxiety because it came in the way that you wake up in the morning and you find messages on WhatsApp showing you must do this and this, causing pressure and tension (**HS, Male, 3, MBchB, 31-35 years)”***.

*“Sometimes you wake and find some messages on twitter, TikTok, watsapp of dead people, or people involved in accidents and your mind goes home immediately, if you happen to call a relative and isn’t picking up or when the phone is off, ahhhhhh…you get worried the whole day. (**FE, Female, 3, WAR, 21-24 years)”***

## Discussion

This study identified social, economic, academic and other contextual factors associated with mental health disorders among students at Busitema University. This was not different from studies carried out at Mbarara University of Science and Technology[10] and other studies which investigated influencing factors of depression among college students in Europe[13]. A detailed analysis of the factors revealed that some of the students were affected by the poor and unconducive learning environments at the faculty of Health sciences compared to the faculty of Engineering which had some infrastructure in place. Regarding the factors read to accommodation, both compasses had similar problems which significantly affected their mental well-being[14]. Family factors, as well as financial factors, were a notch higher than other factors across the two compasses which were similar to a study in Tanzania[15]. Our in-depth analysis revealed that students with financial challenges and broken families experienced psychological distress more often. Another study that was carried out in the United Kingdom indicated that students from well-to-do families were at a lower risk of mental health issues[16]. Exposure to relationships was also a major factor identified by the study which was similar to findings carried out in United kingdom[14]

These findings suggested a specific and feasible approach for the primary prevention of mental health disorders among university students with the utmost involvement of the National Council for Higher Education, the Ministry of Education and Sports as well as key stakeholders from the universities. It is not precisely clear how one goes about promoting resilience; this personality trait may depend on various factors, not all of which can be addressed by an institutional intervention[4]. At the institutional level, there is a call for insight into enhancing resiliency and include support focused on spiritual, emotional and psychological needs as a fundamental prerequisite for students. In other words, students should be taught how to improve their sense of self-worth and to adapt to difficult situations to have a means for effective management of their depressive tendencies[17]. Improvements in self-esteem and self-efficacy would not only strengthen resilience and motivation towards learning and career development, but interventions directed at this goal will also provide internal positive feedback and help students with their performance of difficult tasks while learning to cope in a meaningful and supportive climate[2]. Psychological/ mental well-being among students should be among the highest priorities when developing programs focused on educational interventions.

## Limitations of the study

We prioritized only two campuses because we wanted to get baseline data on what factors were associated with mental health disorders among students at Busitema University.

## Conclusion

Considering the high propensity of mental health issues that hinder the success of students at the university level. It’s worth paramount for universities to continually evaluate the mental health of their students and tailor treatment programs and other cost-effective interventions to specifically target students.

## Data Availability

N/A

## Acknowledgement of the funding body

This study was fully supported by the Busitema University Research and innovation fund grant 3/DGSRI/22. We are grateful to the administration of Busitema University and student leaders of Busitema and Mbale campuses who supported us throughout the data collection process.

## Author contributions

EKK conceived the idea, wrote a proposal with JK and JLM. EKK and JK oversaw and participated data collection, analysis and interpretation of results, drafted manuscript and proofread integrated all co-authors contributions. JLM, SBK, EB, JW, BB, SB, SA, DO and PW supported with proposal writing, data collection, analysis and interpretation of results and proof read all versions of manuscript and provided additional technical support to the team. All authors have read and approved this manuscript prior to submission.

## Data availability statement

The datasets used and analyzed during the current study are available from the corresponding author on reasonable request.

## Competing interests statement

The authors declare no competing interests

## Notes

### Competing Interest Statement

The authors have declared no competing interest.

### Funding Statement

The author received limited funding for this study and the funding was basically to support the data collection exercise.

